# Perceptions and Experiences of Women Using Fertility Tracking Apps: A Protocol for a Systematic Review of Qualitative Studies

**DOI:** 10.1101/2025.06.20.25329976

**Authors:** Ravi Shankar, Fiona Devi, Xu Qian

## Abstract

**Background:** Fertility tracking apps have gained widespread popularity as a convenient method for women to monitor their menstrual cycles and improve chances of conception. Despite their prevalence, there is limited research synthesizing women’s lived experiences and perceptions regarding the use of these apps. Understanding women’s perspectives is crucial to address concerns about accuracy, privacy, and emotional well-being impacts, and to ensure that fertility apps are meeting the needs of their users.

**Objectives:** This systematic review aims to synthesize qualitative research on women’s perceptions and experiences of using fertility tracking apps. The review will explore women’s attitudes, motivations, perceived benefits, and challenges related to the use of these apps. Specifically, it will identify key themes related to how these apps influence women’s emotional well-being, relationships, and decision-making processes, while identifying gaps in current literature to guide future research.

**Methods and Analysis:** This review will consider qualitative studies that include women of reproductive age who have used or are currently using fertility tracking apps. The systematic review will be conducted in accordance with JBI methodology for systematic reviews of qualitative evidence using a meta-aggregative approach. A comprehensive search strategy will be employed across multiple databases, including PubMed, Web of Science, Embase, CINAHL, MEDLINE, The Cochrane Library, PsycINFO, and Scopus, from each database’s inception to January 2025. Two independent reviewers will screen titles and abstracts using Covidence, assess full texts against inclusion criteria, and extract qualitative data using standardized tools. Study quality will be critically appraised using the JBI Critical Appraisal Checklist for Qualitative Research. Data will be analyzed using thematic synthesis following a three-step meta-aggregative approach: extracting findings verbatim, grouping similar findings into categories, and synthesizing categories into overarching findings. The certainty of findings will be assessed using the ConQual approach.

**Discussion:** This systematic review will provide the first comprehensive synthesis of women’s experiences with fertility tracking apps, generating actionable insights for app developers, healthcare providers, and researchers. The synthesized findings can inform the development and refinement of fertility tracking apps to better meet user needs and preferences. Healthcare providers may benefit from these insights to guide patient discussions about fertility tracking and provide more tailored support. The identification of literature gaps will help direct future research efforts toward more diverse and inclusive studies on fertility tracking apps, ultimately contributing to improved women’s reproductive health and well-being.

**PROSPERO registration number:** CRD42025645316

## Introduction

Fertility tracking apps, also known as period tracking or ovulation tracking apps, have emerged as popular tools for women to monitor their menstrual cycles and optimize their chances of conceiving [1]. These apps allow users to log various data points, such as menstrual dates, basal body temperature, and cervical mucus consistency, to predict ovulation and identify the fertile window [2]. Some apps also offer additional features like community forums, educational resources, and integration with wearable devices [3].

The use of fertility tracking apps has increased significantly in recent years. A 2019 survey found that nearly one-third of women in the United States had used a period or fertility tracking app at some point [4]. The global mobile health (mHealth) app market, which includes fertility apps, is projected to reach $111.1 billion by 2025 [5]. This growing prevalence underscores the need to understand women’s experiences and perceptions surrounding these apps.

While fertility tracking apps offer potential benefits, such as increased awareness of fertility signs and a sense of control over reproductive health, concerns have also been raised. Accuracy of predictions, data privacy and security, and the potential impact on emotional well-being are among the issues that have been highlighted [6,7]. A deeper understanding of women’s perspectives is crucial to address these concerns and ensure that fertility apps are meeting the needs of their users.

Qualitative research provides valuable insights into individuals’ lived experiences, beliefs, and decision-making processes. Several qualitative studies have explored women’s experiences with fertility tracking apps, revealing themes such as empowerment, anxiety, and the impact on relationships [8,9]. However, there has been no comprehensive synthesis of these findings to date.

A systematic review of qualitative studies on women’s experiences with fertility tracking apps would provide a consolidated understanding of the current state of knowledge in this area. By synthesizing findings across multiple studies, this review can identify overarching themes, gaps in the literature, and areas for future research. The insights gained from this review could inform the development and refinement of fertility tracking apps, as well as healthcare practices related to fertility management.

The proposed systematic review will be guided by the Joanna Briggs Institute (JBI) methodology for qualitative systematic reviews [10]. The JBI approach is based on the meta-aggregative method, which involves the synthesis of findings from qualitative studies to produce generalized statements in the form of recommendations that can guide practitioners [11]. This approach is particularly suitable for the current review, as it allows for the aggregation of findings across multiple qualitative studies to generate actionable insights for app developers, healthcare providers, and researchers.

This systematic review seeks to address the question: What are women’s perceptions and experiences of using fertility tracking apps? The objective is to synthesize existing qualitative research to gain a deeper understanding of how women engage with these digital tools. Specifically, the review aims to identify and summarize key themes related to women’s motivations, attitudes, and experiences with fertility tracking apps. It will explore the perceived benefits and challenges associated with their use and examine how these apps influence women’s emotional well-being, relationships, and decision-making processes. Furthermore, the review will identify gaps in the current body of literature and provide recommendations to guide future research in this area.

## Methods

The proposed systematic review will be conducted in accordance with the JBI methodology for systematic reviews of qualitative evidence [12].

### Inclusion criteria Types of participants

This review will consider studies that include women of reproductive age (generally 15-49 years old) who have used or are currently using fertility tracking apps. Studies involving healthcare providers or male partners of app users will be excluded unless they also include the perspectives of female app users.

### Phenomena of interest

The phenomena of interest are women’s perceptions, attitudes, and experiences related to the use of fertility tracking apps. This includes, but is not limited to, their motivations for using these apps, perceived benefits and challenges, the impact on their emotional well-being, relationships, and decision-making processes, as well as their overall satisfaction with the apps.

### Context

The context of this review is global, as fertility tracking apps are used worldwide. However, the review will only include studies published in English due to resource constraints.

### Types of studies

This review will consider qualitative studies, including, but not limited to, designs such as phenomenology, grounded theory, ethnography, action research, and feminist research. Mixed-methods studies will be considered if the qualitative component is clearly identifiable and extractable. Quantitative studies, editorials, opinion pieces, and narrative reviews will be excluded.

### Search strategy

The search strategy will aim to locate both published and unpublished studies. An initial limited search of PubMed and CINAHL will be undertaken to identify articles on the topic. The text words contained in the titles and abstracts of relevant articles, and the index terms used to describe the articles, will be used to develop a full search strategy for each database (see Appendix I). The search strategy, including all identified keywords and index terms, will be adapted for each included database and/or information source. The reference lists of all included studies will be screened for additional papers.

The literature search for this systematic review will be conducted across several electronic databases to ensure comprehensive coverage of relevant studies. These databases include PubMed, Web of Science, Embase, CINAHL, MEDLINE, The Cochrane Library, PsycINFO, and Scopus. In addition to published literature, sources of unpublished studies and gray literature will also be explored to minimize publication bias. These sources include ProQuest Dissertations and Theses, OpenGrey, and Google Scholar.

### Study selection

Following the search, all identified records will be collated and uploaded into Covidence systematic review software [13] and duplicates removed. Titles and abstracts will then be screened by two independent reviewers for assessment against the inclusion criteria for the review. Potentially relevant papers will be retrieved in full and their citation details imported into Covidence. The full text of selected citations will then be assessed in detail against the inclusion criteria by two independent reviewers. Reasons for exclusion of papers at full text that do not meet the inclusion criteria will be recorded and reported in the systematic review. Any disagreements that arise between the reviewers at each stage of the selection process will be resolved through discussion or with a third reviewer. The results of the search and the study inclusion process will be reported in full in the final systematic review and presented in a PRISMA flow diagram [14].

### Assessment of methodological quality

Qualitative papers selected for retrieval will be assessed by two independent reviewers for methodological quality using the JBI Critical Appraisal Checklist for Qualitative Research [15]. Any disagreements that arise between the reviewers will be resolved through discussion or with a third reviewer. All studies, regardless of the results of their methodological quality, will undergo data extraction and synthesis (where possible).

The JBI Critical Appraisal Checklist for Qualitative Research comprises ten questions that assess various aspects of methodological quality [15]. These questions are designed to evaluate the congruence and coherence within a qualitative study. Specifically, the checklist examines whether there is alignment between the stated philosophical perspective and the research methodology, as well as between the methodology and the research question or objectives. It also considers the consistency between the methodology and the methods used to collect data, the representation and analysis of data, and the interpretation of results. Additionally, the checklist looks for a statement that locates the researcher culturally or theoretically and assesses whether the influence of the researcher on the research (and vice versa) is addressed. It also evaluates whether participants and their voices are adequately represented, whether the study meets ethical standards or provides evidence of ethical approval, and whether the conclusions are supported by the data analysis or interpretation.

Each question is answered with “yes,” “no,” “unclear,” or “not applicable.” A “yes” response indicates that the study meets the criteria for that particular question. The overall appraisal of each study will be based on the number of “yes” responses, with a higher number indicating a higher level of methodological quality.

### Data extraction

Qualitative data will be extracted from papers included in the review by two independent reviewers using the standardized data extraction tool in Covidence [13]. The data extracted will include specific details about the populations, context, culture, geographical location, study methods, and the phenomena of interest relevant to the review question and objectives. Findings and their illustrations will be extracted and assigned a level of credibility.

The data extraction form in Covidence includes several key fields designed to capture comprehensive information from each study. These fields include study details (such as authors, year of publication, country, and study design), participant characteristics (including the number of participants, age, fertility status, and app usage), and app characteristics (such as the app name, features, and platform). In addition, details about study methods (data collection and analysis methods) and study findings (themes, subthemes, supporting quotes, and interpretations) are extracted. A section for the reviewer’s notes is also included to capture any relevant observations or comments. During the data extraction process, reviewers will assign a level of credibility to each finding based on the JBI Levels of Credibility. These levels are categorized as: *unequivocal*—findings supported by evidence beyond reasonable doubt and not open to challenge; *credible*—findings that are supported by interpretive illustrations and open to challenge; and *unsupported*—findings lacking data support. This assessment of credibility will play a critical role in guiding the data synthesis process and shaping the level of confidence in the final synthesized findings.

### Data synthesis

Qualitative research findings will be pooled using JBI methodology. This will involve the aggregation or synthesis of findings to generate a set of statements that represent that aggregation, through assembling the findings and categorizing these findings based on similarity in meaning. These categories will then be subjected to a meta-synthesis in order to produce a comprehensive set of synthesized findings that can be used as a basis for evidence-based practice. Where textual pooling is not possible, the findings will be presented in narrative form.

The data synthesis process for this review will follow the three-step approach outlined by Lockwood et al. [12]. First, all findings from the included studies, along with their accompanying illustrations, will be extracted verbatim. Each finding will then be assessed and assigned a level of credibility—unequivocal, credible, or unsupported— based on the JBI criteria. In the second step, findings that are sufficiently similar in meaning will be grouped into categories. This involves a thorough and iterative reading of the findings to identify patterns and commonalities. Finally, in the third step, two or more categories will be synthesized to generate overarching synthesized findings. These synthesized findings will aim to provide a comprehensive understanding of the topic and serve as a foundation for evidence-based practice.

Throughout the data synthesis process, the reviewers will consider the levels of credibility assigned to each finding during data extraction. Findings with higher levels of credibility (i.e., unequivocal and credible) will be given more weight in the synthesis compared to findings with lower levels of credibility (i.e., unsupported) [16].

The synthesized findings will be presented narratively, along with a summary of the categories and findings that contributed to each synthesized finding. Relevant quotes from the included studies will be used to illustrate the synthesized findings.

### Assessing the certainty of the findings

The final synthesized findings of this review will be graded using the ConQual approach, which is designed to establish the level of confidence in qualitative research synthesis.

ConQual assesses two key components: *dependability* and *credibility* of each synthesized finding. Dependability is evaluated based on several criteria, including the appropriateness of the study design and methods for addressing the research question, the congruence between the research methodology and the methods used for data collection, analysis, and interpretation, the degree to which the researchers’ influence on the findings is acknowledged, and the extent to which participants and their voices are adequately represented in the findings. Credibility, on the other hand, is assessed by examining the alignment between the data from the primary studies and the synthesized findings, as well as the levels of credibility (unequivocal, credible, or unsupported) assigned to individual findings during the data extraction process [17]. Together, these assessments contribute to a transparent and structured evaluation of the confidence that can be placed in the synthesized qualitative evidence.

Based on the assessment of dependability and credibility, each synthesized finding will be assigned a ConQual score, ranging from “high” to “very low.” The ConQual scores will be presented in a Summary of Findings table, along with an explanation of the reasons for each score.

## Discussion

This protocol outlines a systematic review of qualitative studies that aims to synthesize women’s perceptions and experiences of using fertility tracking apps. The review will be guided by the JBI methodology for qualitative systematic reviews and will adhere to the PRISMA guidelines for reporting systematic reviews.

The strengths of this review include its comprehensive search strategy, which covers multiple databases and sources of gray literature, as well as its rigorous methodological approach. The use of two independent reviewers for study selection, critical appraisal, and data extraction will minimize the risk of bias and ensure the reliability of the findings. The application of the JBI meta-aggregative approach to data synthesis will allow for the generation of actionable recommendations for practice based on the synthesized findings.

However, there are also some potential limitations to consider. The restriction to studies published in English may exclude relevant research conducted in other languages. The inclusion of only qualitative studies may limit the generalizability of the findings to broader populations. Additionally, the rapidly evolving nature of mobile apps means that some of the findings may not be applicable to newer fertility tracking apps with different features and functionalities.

Despite these limitations, this systematic review has the potential to make a significant contribution to the understanding of women’s experiences with fertility tracking apps. The synthesized findings can inform the development and refinement of these apps to better meet the needs and preferences of their users. Healthcare providers may also benefit from the insights gained from this review, as they can use the findings to guide their discussions with patients about fertility tracking and provide more tailored support.

Furthermore, the identification of gaps in the current literature can help direct future research efforts in this area. For example, if the review reveals a lack of studies focusing on certain populations or geographical regions, this can highlight the need for more diverse and inclusive research on fertility tracking apps.

In conclusion, this systematic review protocol presents a rigorous and comprehensive approach to synthesizing qualitative research on women’s perceptions and experiences of using fertility tracking apps. The findings of this review have the potential to inform app development, healthcare practices, and future research, ultimately contributing to the improvement of women’s reproductive health and well-being.

## Data availability

Data supporting the findings is available upon request. Please contact the Corresponding Author, Dr Ravi Shankar for data availability.

## Appendix I Search strategy PubMed

(“Fertility Tracking”[All Fields] OR “Period Tracking”[All Fields] OR “Ovulation Tracking”[All Fields] OR “Menstrual Cycle Tracking”[All Fields]) AND (“Mobile Applications”[MeSH Terms] OR “mobile app*”[All Fields] OR “smartphone app*”[All Fields]) AND (“Perception”[MeSH Terms] OR “perception*”[All Fields] OR “Attitude”[MeSH Terms] OR “attitude*”[All Fields] OR “Experience*”[All Fields] OR “Motivation*”[All Fields] OR “Challenge*”[All Fields] OR “Benefit*”[All Fields] OR “Empower*”[All Fields] OR “Anxiety”[MeSH Terms] OR “anxi*”[All Fields] OR “relationship*”[All Fields] OR “well-being”[All Fields] OR “Decision Making”[MeSH Terms] OR “decision making”[All Fields]) AND (“Qualitative Research”[MeSH Terms] OR “qualitative”[All Fields] OR “Interview*”[All Fields] OR “Focus Groups”[MeSH Terms] OR “focus group*”[All Fields] OR “phenomenolog*”[All Fields] OR “grounded theory”[All Fields] OR “ethnograph*”[All Fields] OR “action research”[All Fields] OR “feminist”[All Fields] OR “mixed method*”[All Fields])

## Appendix II Data Extraction Form

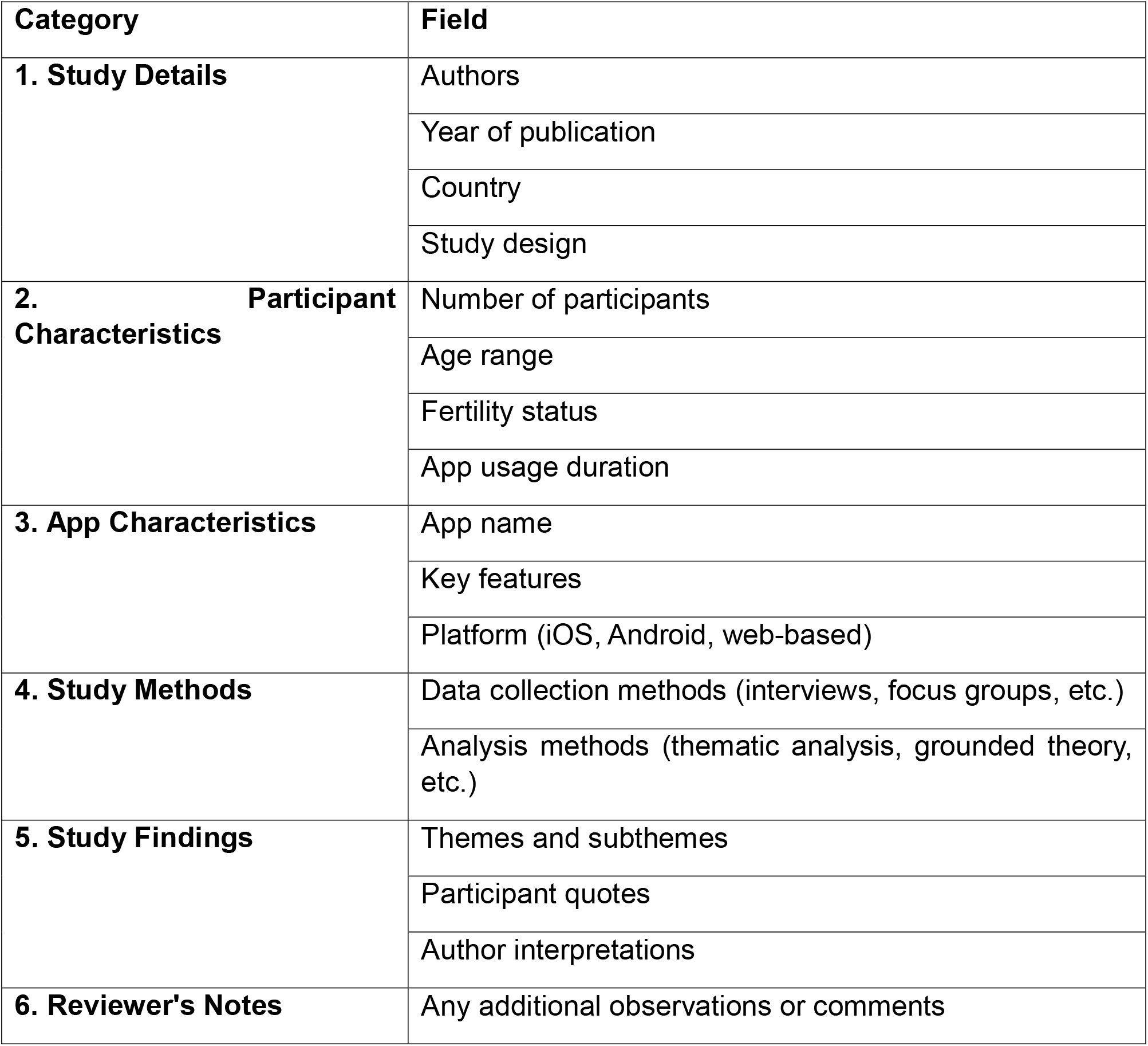

Appendix II: Data Extraction Form
| Category | Field |
| --- | --- |
| **1. Study Details** | Authors |
|  | Year of publication |
|  | Country |
|  | Study design |
| **2. Participant Characteristics** | Number of participants |
|  | Age range |
|  | Fertility status |
|  | App usage duration |
| **3. App Characteristics** | App name |
|  | Key features |
|  | Platform (iOS, Android, web-based) |
| **4. Study Methods** | Data collection methods (interviews, focus groups, etc.) |
|  | Analysis methods (thematic analysis, grounded theory, etc.) |
| **5. Study Findings** | Themes and subthemes |
|  | Participant quotes |
|  | Author interpretations |
| **6. Reviewer's Notes** | Any additional observations or comments |

